# Risk Ratio (Relative Risk) And Odds Ratio For Radiologically Visible Intracranial Air After Neuraxial Access Attempts Among Our Peri-Partum Patients Were Undefined Although One In Forty Of Them Had Radiologically Visible Intracranial Air On CT/MR Of Head/Brain

**DOI:** 10.1101/2022.10.02.22280605

**Authors:** Deepak Gupta

**Author notes:** **Corresponding Author:** Dr Deepak Gupta, Clinical Assistant Professor, Anesthesiology, Wayne State University/Detroit Medical Center, Box No 162, 3990 John R, Detroit, MI 48201, United States.

## Abstract

**Background:** During neuraxial access attempts, loss-of-resistance technique with air may inadvertently become injection-of-air technique potentially leading to intracranial air.

**Objective:** The purpose of this retrospective study was to ascertain the incidence of radiologically visible intracranial air among peri-partum patients admitted to local Women’s Hospital who underwent computed tomography/magnetic resonance imaging (CT/MR) of head/brain during their hospital stay at peri-partum floors over a seven-year period of July 1, 2015-June 30, 2022.

**Methods:** After Institutional Review Board approval for exempt research, medical records of patients who underwent CT/MR of head/brain during their hospital stay at peri-partum floors over a seven-year period of July 1, 2015-June 30, 2022 were reviewed to see whether they had neuraxial access attempts (epidurals, spinals, combined spinal-epidurals, epidural blood patches) before their CT/MR of head/brain. Subsequently, such patients CT/MR of head/brain were reviewed to ascertain the evidence of any radiologically visible intracranial air.

**Results:** Only 69 peri-partum patients underwent CT/MR of head/brain during the seven-year period with 40 of them receiving neuraxial access attempts before their CT/MR of head/brain. Only one labor epidural analgesia patient had radiologically visible intracranial air.

**Conclusion:** In seven-year period sample among our peri-partum patients at local Women’s Hospital, risk ratio (relative risk) and odds ratio for radiologically visible intracranial air after neuraxial access attempts were undefined although one in forty had radiologically visible intracranial air on CT/MR of head/brain after neuraxial access attempts and that intracranial air was 100% attributable to neuraxial access attempts.

## Introduction

This retrospective study’s question was to determine the local institutional incidence of radiologically evident presence of intracranial air (pneumocephalus) in peri-partum patients considering that many if not all peri-partum patients undergo neuraxial access attempts for labor analgesia, surgical anesthesia and post dural puncture headache managements in the form of epidurals, combined spinal-epidurals and epidural blood patches respectively wherein loss-of-resistance technique with air is most commonly utilized that can inadvertently inject air into epidural space that may indirectly travel into intrtathecal space unless inadvertent dural puncture during epidural access directly instills air into intrathecal space just like persistent air bubbles within the medication syringe while medication is being injected intrathecally during administration of spinal anesthesia [1-12].

The purpose of this retrospective study was to ascertain the incidence of radiologically visible intracranial air among peri-partum patients admitted to local Women’s Hospital who underwent computed tomography/magnetic resonance imaging (CT/MR) of head/brain during their hospital stay at peri-partum floors over a seven-year period of July 1, 2015-June 30, 2022.

## Methods

After Institutional Review Board approval for exempt research, our institute’s information technology team ran a query in institutional electronic medical records’ database to extract local Women’s Hospital’s peri-partum patients’ list for July 1, 2015-June 30, 2022 (seven-year period) who had undergone CT/MR of head/brain during their hospital stay. Thereafter, such patients’ medical records were reviewed to see whether they had neuraxial access attempts (epidurals, spinals, combined spinal-epidurals, epidural blood patches) before their CT/MR of head/brain. Subsequently, such patients CT/MR of head/brain were reviewed to ascertain the evidence of any radiologically visible intracranial air. Finally, per our anesthesia billing department, the total number of peri-partum patients billed for anesthesia care including but not limited to neuraxial access attempts (epidurals, spinals, combined spinal-epidurals and epidural blood patches) were ascertained so as to put the above-mentioned sample’s data into perspective for a true estimate of intracranial air incidence among peri-partum patients who were concurrently our peri-anesthesia patients too.

## Statistical Analysis

The data was compared by 2×2-contingency table as per given below wherein as per detailed instructions [13], relative risk (risk ratio) for intracranial air with neuraxial access attempts was deciphered as {A/(A+B)}/{C/(C+D)}; odds ratio was deciphered as (A/B)/(C/D); and attributable risk was deciphered as {A/(A+B)}-{C/(C+D)}:

|  | Number of patients<br>who had<br>radiologically<br>visible intracranial<br>air | Number of patients<br>who did NOT have<br>radiologically<br>visible intracranial<br>air | TOTAL |
| --- | --- | --- | --- |
| Number of patients<br>who had neuraxial<br>access attempts<br>before radiological<br>exams | A | B | A+B |
| Number of patients<br>who did NOT have<br>neuraxial access<br>attempts before<br>radiological exams | C | D | C+D |
| TOTAL | A+C | B+D | A+B+C+D |

## Results

During the seven-year period, only 132 patients had CT/MR of head/brain during their hospital stay at peri-partum floors in local Women’s Hospital. However, only 69 patients among these 132 patients were truly peri-partum who had delivered vaginally or via cesarean section before or after CT/MR of head/brain during their hospital admissions. Among these 69 peri-partum patients, 29 patients did not have any recorded neuraxial access attempts before CT/MR of head/brain. In contrast, 28 patients had epidural access attempts for labor analgesia before CT/MR of head/brain as well as 12 patients had spinal access attempts for surgical anesthesia before CT/MR of head/brain. Additionally, three out of 28 labor epidural analgesia patients had epidural blood patch access attempts before CT/MR of head/brain, while two out of 12 spinal anesthesia patients had epidural blood patch access attempts before CT/MR of head/brain. Essentially, a total of 30 peri-partum patients had epidural space access attempts with loss-of-resistance technique before CT/MR of head/brain while a total of 10 peri-partum patients had direct intrathecal space access attempts without loss-of-resistance technique before CT/MR of head/brain. Among all these patients, only one labor epidural analgesia patient had radiologically visible intracranial air.

Therefore, the statistics data table became as follows:

|  | Number of patients<br>who had<br>radiologically<br>visible intracranial<br>air | Number of patients<br>who did NOT have<br>radiologically<br>visible intracranial<br>air | TOTAL |
| --- | --- | --- | --- |
| Number of patients<br>who had neuraxial<br>access attempts<br>before radiological<br>exams | 1 | 39 | 40 |
| Number of patients<br>who did NOT have<br>neuraxial access<br>attempts before<br>radiological exams | 0 | 29 | 29 |
| TOTAL | 1 | 68 | 69 |

Consequently, relative risk (risk ratio) for intracranial air with neuraxial access attempts was deciphered as (1/40)/(0/29)=Undefined; odds ratio was deciphered as (1/39)/(0/29)=Undefined; and attributable risk was deciphered as (1/40)-(0/29)=0.025 with both attributable risk percentage as well as population attributable risk percentage being 100% [14-15]. In a nutshell, although risk ratio and odds ratio in the current sample of data could not be defined due to division by zero, the attributable risk percentages suggested that 100% of radiologically visible intracranial air among peri-partum patients was attributable to neuraxial access attempts among them before their radiological examinations. Interestingly, the surprisingly miniscule sample size of 69 patients data over seven-year period negated any possibility of extrapolating to any true estimate of intracranial air incidence among peri-partum patients during the same seven-year period who were concurrently our peri-anesthesia patients too with a total of 88 epidural blood patches billed by our anesthesia department during the same seven-year period after having billed anesthesia care for a total of 14,947 labor epidural analgesia patients plus a total of 5,730 cesarean section patients under neuraxial and/or general anesthesia during the same seven-year period.

## Discussion

The only key finding in the current miniscule sample of 69 peri-partum patients’ data over a seven-year period was that one out of forty (2.5%) neuraxially accessed peri-partum patients with one out of thirty (3.3%) epidurally accessed peri-partum patients had radiologically visible intracranial air with 100% of radiologically visible intracranial air among peri-partum patients attributable to neuraxial (and epidural) access attempts among them before their radiological examinations because more often than not, loss-of-resistance technique with air during epidural access attempts inadvertently becomes injection-of-air technique as observed anecdotally [16].

The current study has some limitations. Firstly, the sample data size turned out to be minuscule unless it may be indicating that truly real percentage of peri-partum patients receiving peri-anesthesia CT/MR of head/brain at local Women’s Hospital was <1% {69*100/(14,947+5,730)}. Secondly, the sample data could have further expanded its explorative reach by including CT/MR of neck/cervical spine because extremely limited incidence of intracranial air deciphered in the current sample data might have concretely and statistically expanded with inclusion of radiologically visible intraspinal air among peri-partum patients especially when pneumocephalus and pneumorrachis may coexist to present as potentially concurrent headache and neckache after neuraxial (especially epidural) access attempts among peri-partum patients.

## Conclusion

In a seven-year period sample among our peri-partum patients at local Women’s Hospital, risk ratio (relative risk) and odds ratio for radiologically visible intracranial air after neuraxial access attempts were undefined although one in forty had radiologically visible intracranial air on CT/MR of head/brain after neuraxial access attempts and that intracranial air was 100% attributable to neuraxial access attempts.

## Supporting information

INSTITUTIONAL BOARD APPROVAL

## Data Availability

All data produced in the present work are contained in the manuscript

