## Supplementary material for "Risk Ratio (Relative Risk) And Odds Ratio For Radiologically Visible Intracranial Air After Neuraxial Access Attempts Among Our Peri-Partum Patients Were Undefined Although One In Forty Of Them Had Radiologically Visible Intracranial Air On CT/MR Of Head/Brain": INSTITUTIONAL BOARD APPROVAL

WAYNE STATE UNIVERSITY

IRB Administration Office
87 East Canfield, Second Floor
Detroit, MI 48201
www.irb.wayne.edu

### CONCURRENCE OF EXEMPTION
#### IRB-22-07-4792-MP2 Expedited/Exempt-EXEMPT

| | |
|---|---|
| **DATE:** | July 28, 2022 |
| **TO:** | Gupta, Deepak, Anesthesiology<br>Zestos, Maria, Deans Office Medicine |
| **FROM:** | Smitherman, Lynn, Associate Professor - Clinical, MP2 Expedited/Exempt |
| **PROTOCOL TITLE:** | How Common Is Intracranial Air (Pneumocephalus) In Peri-partum Patients? |
| **FUNDING SOURCE:** | None |
| **PROTOCOL NUMBER:** | IRB-22-07-4792<br>Approval Date: July 28, 2022 |

The above-referenced protocol has been reviewed and found to qualify for Exemption according to category 4

**Note to PI:** This IRB approval does not replace administrative or department/college/division approvals that may be required. Before initiating research activities contact the Associate/Vice Dean for Research in your school or college for established parameters for site access to the facility where the study will be conducted.

**NOTE TO PRINCIPAL INVESTIGATORS**: Due to the COVID-19 health crisis, the resumption of human participant research is occurring in measured phases which incorporate institutional, state, and federal regulations and best practices.

Currently the following research activities are ongoing:

(I) Human participant research that can maintain remote study interventions/visits as per IRB approval.

(II) In-person research with a direct benefit and in-person research that has no potential for direct benefit conducted at Wayne State University and/or established health care facilities.

(III) In-person research without potential for direct benefit to participants conducted at non-affiliated WSU sites must include the non-affiliated study sites' approval/letter of support to conduct in-person research activities.

(IV) **Effective September 1, 2021:** For research conducted at WSU campus sites: Participants must complete the WSU Campus Guest Screener. The WSU campus vaccine mandate is not required for research participants who are visiting campus to participate in a research study. However, mitigation plans as indicated for Appendix N must be followed. The research participant must be contacted before the study visit to inform them of the WSU campus screening and safety precautions.

Information on restarting WSU research operations can be found at research.wayne.edu/irb/coronavirus

In-person research activities require additional precautions to protect both the participant and the research staff. Mitigation procedures indicated for Appendix N must be followed.

For more information regarding Appendix N and IRB resumption of research requirements visit: research.wayne.edu/irb/coronavirus.

When clinical research is conducted in a standard medical care/hospital setting, please follow that site ’s COVID-19 precautionary standard operating procedures Appendix N is not required.

For more information regarding IRB submission requirements and instructions visit the IRB Forms and Submissions Requirements website: research.wayne.edu/irb/forms-requirements-categories.

If you have questions please contact the IRB Administration Office, or telephone: 313-577-1628.

IRB Administration Office
87 East Canfield, Second Floor
Detroit, MI 48201
www.irb.wayne.edu

The following attachments and consent/assent documents have been reviewed and approved by the IRB.

Notes:
NOTE TO PI: This project has been given a Status Check-In Date. The Status Check-In Date is 07/28/2024. The Minimal Risk Status Update Form should be used to provide a status report to the IRB. Please submit the status update at least 6 weeks before this date. The Minimal Risk Status Update Form is available on the IRB's Forms and Submissions website (www.irb.wayne.edu). The Minimal Risk Status Update should be submitted as an expedited amendment via eProtocol with the Minimal Risk Status Update Form. Include the Minimal Risk Status Update Form as an Attachment using the label: Minimal Risk Status Update.

Protocol/Proposal/Dissertation (dated 07/2022)

The following data collection materials have been reviewed and approved and does not require a WSU IRB stamp for use. These documents are approved and noted in the IRB file (1): Data Collection Sheet.

A waiver of consent has been granted according to 45CFR46.116(d). This waiver satisfies: 1) risk is no more than minimal, 2) the waiver does not adversely affect the rights and welfare of research participants, 3) the research could not be practicably carried out without the waiver, and 4) the participants will not be given information.

A waiver of HIPAA Authorization has been granted in accordance with the Privacy Rule and Justification provided by the Principal Investigator in the HIPAA Summary Form. This waiver satisfies: 1) the use or closure of PHI involves no more than minimal risk to the privacy of individuals, 2) the research could not be practicably conducted without the waiver, 3) the research could not be practicably conducted without access and use of the PHI, 4) adequate steps taken to protect identifiers from improper use of disclosure and 5) adequate plan for destroying identifiers or links.

* Exempt protocols do not require annual review by the IRB, however you may have been granted a Status Check-In Date. Projects granted a Status Check-In date must submit a Minimal Risk Status Update Report at least 6 weeks before the check-In date. If research activities are complete a Final Report/Closure must be submitted by the Status Check-In date.

* All changes or amendments to the above-referenced protocol require review and approval by the IRB BEFORE implementation.

* Adverse Reactions/Unanticipated Problems AR/UP must be submitted on the appropriate form within the time frame specified in the IRB. In the event of an unanticipated problem use the Unanticipated Problem Report Form located on the IRB's Forms and Submissions Requirements website.

Note: Studies conducted at DMC sites or DMC medical record used for affiliate review Authorized DMC personnel have been added to this submission under Personnel Information "Other".
Administration Office Policy www.irb.wayne.edu/policies-human-research

NOTE: Upon notification of an impending regulatory site visit, hold notification, and/or external audit the IRB Administration Office must be contacted immediately. Also Notify the IRB of any changes to the funding status of the above-referenced protocol.

**Attachments**

- CV Deepak Gupta 2022
- Research Review Process
- DATA WORKSHEET
- Protocol UPDATED
- Protocol

WAYNE STATE UNIVERSITY

IRB Administration Office
87 East Canfield, Second Floor
Detroit, MI 48201
www.irb.wayne.edu

---

| | |
|---|---|
| **Review Type:** | EXEMPT |
| **IRB Number:** | MP2 Expedited/Exempt Review |
